# Review of spatial planning policy as an intervention for improving public health: protocol for a health-focused content review of local plans in England

**DOI:** 10.1101/2023.06.22.23291736

**Authors:** Michael Chang, Neil Carhart

## Abstract

**Introduction:** Each local authority in England is required to create a statutory local plan that sets out their land use priorities as the basis of decision-making on urban development. Only recently has there been an explicit recognition the local plan is capable of being a legal determinant of and important public policy intervention in influencing population physical and mental health and wellbeing. But there is a lack of comprehensive understanding of whether and how local plans have integrated wider determinants of health. The objective of this research is to identify and understand the state of current local plans in relation to health by undertaking a census.

**Methods and analysis:** The review for England will involve a census of all local plans, and content and thematic analyses of a sample of local plans. This is subject to developing a review framework as the basis for gathering findings using a combination of query functions in PDF documents and data presentation in Excel, supported latterly by the computerassisted qualitative data analysis software, NVivo. The review framework will include a range of search parameters for identifying explicit health references across specified policy topics and wider determinants of health indicators as they relate to spatial planning based on an established evidence publication. A sampling strategy identifies inclusion and exclusion criteria to focus on local plans in England used in planning decision making. Results will be collected in qualitative and quantitative data formats. The results will be synthesised and discussed using a combination of qualitative data analytical strategies.

**Ethics and dissemination:** The review is desktop based using publicly available policy documents on local authority websites, so ethics approval is not required. The results of the review will be reported in journal articles and for policy makers to improve national and local policy development.

## 1. Introduction

In planning legislation, each local authority is required to have local plans that “*set out the authority’s policies relating to the development and use of land in their area*” (Planning and Compulsory Purchase Act 2004). The local plan is developed through a process of evidence gathering, consultation, independent testing by government inspectors then formally adopted by the local planning authority (LPA), which puts it into effect. The local plan should be reviewed every five years to keep it up to date but it is used and still in effect until a time it is superseded by a more up to date adopted local plan.

These land use requirements for local plans are often set out by national governments through expression in national policy frameworks. In England this is the National Planning Policy Framework (the NPPF)[1]. The composition and content of the local plan is complex and mandated through a combination of primary legislation, secondary regulations, national policy and national guidance, then operationalised in an evidence-based way through the local plan-making process. While in law the local plan as a policy instrument has not changed fundamentally since the Planning and Compulsory Purchase Act 2004, changes in national political priorities and policy framing have evolved especially after the transformational shift in political landscape in 2010 with the Coalition Government replacing the Labour Government. Planning reform programme from 2020 as the result of the Planning White Paper is expected to introduce further changes in national policy as well as the scope and purpose of local plans.

The local plan is a critical component of the spatial planning process in relation to improving health and wellbeing [2]. It is strategically timely as planning reform in England sets out proposals to reform the local planning process and aims to simplify the content of local plans [3]. Having an understanding of how current local plans are structured and what policies they contain can provide insight on how they can be improved upon to address health, and also how the proposed introduction of national planning policies can provide consistency across all LPAs.

In relation to planning for health in England, the NPPF sets out the following strategic directives for local plan policies to[1]:

- enable and support healthy lifestyles, especially where this would address identified local health and well-being needs
- take into account and support the delivery of local strategies to improve health, social and cultural well-being for all sections of the community.

One approach to analysis is to identify local plan policy compliance with the above directives. But policy compliance alone will not be able to determine whether health outcomes are considered across the whole local plan. This is because the NPPF, as a public policy document, is subject to wider political and professional balance, such as political acceptability on certain subject matters than purely informed by evidence. In addition, as a public policy document, it is important to distinguish policy responsibilities across UK government departments with planning responsibilities residing in the Department for Levelling Up, Housing and Communities while health responsibilities residing in the Department of Health and Social Care. This discourse about spatial planning for health through the public policy lens provides an interesting juxtaposition between rational and evidence-informed policy and practice [4], which is grounded in public health practice versus what Thomas Dye refers to as “public policy is whatever governments choose to do or not to do”[5].

In addition, the NPPF also sets out a health across other policy areas that broadly aligns with the Spatial Planning and Health evidence review[6]. So the basis of a review assessment framework emerges for this research on the local plan. Office for Health Improvement and Disparities^1^ is beginning to articulate guidance to local practitioners on this framework to ensure policy compliance but that can also achieve health outcomes from research[6-8] (see Figure 1).

**Figure 1:**
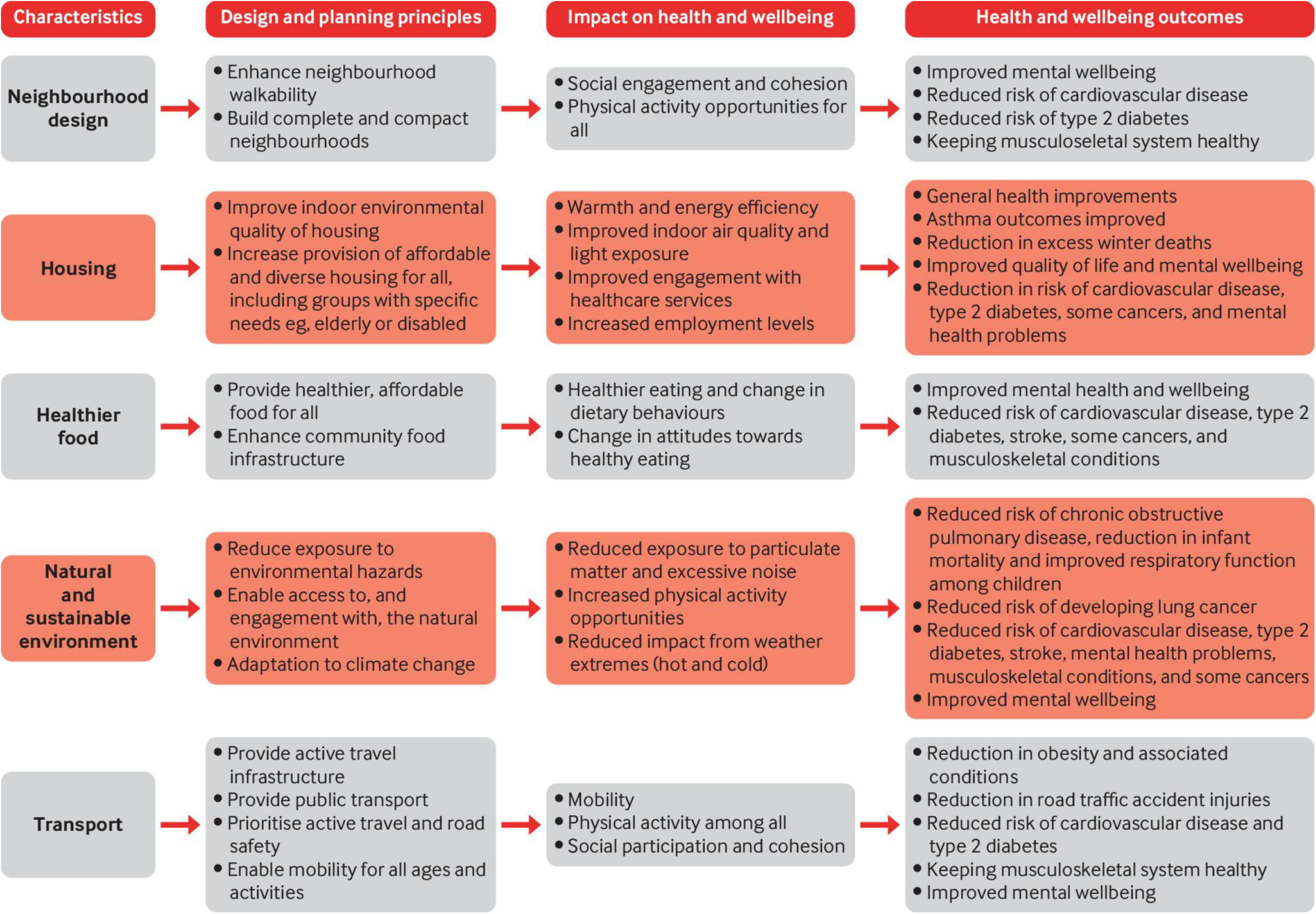
Public Health England framework mapping evidence review against national planning policy requirements.

There are recent local plans review in England which will act as useful reference points in future analysis. The most relevant ones include a health-focused content review of local plans in England and Wales conducted by the Town and Country Planning Association in 2018 as part of its review into the state of the union between spatial planning and public health[9] and a health-focused sample comparative review of local plans in England in 2022 as a specific study to inform a local authority’s preparation of their local plan review process[10]. Otherwise undertaking policy reviews of local plans for health has been limited apart those reviews with a specific purpose to find whether something exists or not, such as whether there are policies on managing hot food takeaways[11] or on older people housing provision[12]. As such, the majority of such local plan reviews have been undertaken outside academia as grey literature often by private sector consultancies and third sector groups to advocate for a certain policy agenda.

There are several potential reasons why comprehensive policy reviews, or census, of local plans have not been subject to extensive undertaking, including the scale of the sample size of more than 300 local plans that makes the task particularly resource-intensive, and limited recognition and understanding within the research sector of respective public health and planning systems that makes initiating such trans-disciplinary research challenging[13]. Local plans are also synonymous as the statutory development plan for the local authority which will comprise of more than one singular document that may not all be created then adopted at the same time. There is no template or consistent format, content or style mandated by government and there is no stipulation of the complete range of subject matters local plan must include if already articulated in national policy and guidance. Therefore all local plans vary in size, scope and detail which is one of the reasons why how health is or is not articulated in local plans continues to be a chronic barrier in delivering health outcomes through spatial planning and urban development decisions[14-16].

But these challenges do not diminish the need to build a novel census of what currently exists in local plans on health. This baseline and reference point of the state of current policy is necessary if we are to begin to systematically and effectively, informed by evidence, make in-roads to improving health, wellbeing and equity through spatial planning.

## 2. Study objective and outcomes

The local plan policy review aims to address a hypothesis within the overall research that health in spatial planning policies is a necessary prerequisite to achieving healthy places. This hypothesis will be tested by this review to gain a foundational understanding and identifying what wider determinants of health indicators are contained in planning policies within each local authority’s local plan in England. It will then inform further systems research activities with selected local authorities to better understand how health has been embedded in their local plan processes and relationships. An early logic model was created as part of establishing this protocol (see Figure 2). The following outcomes are expected:

**Figure 2:**
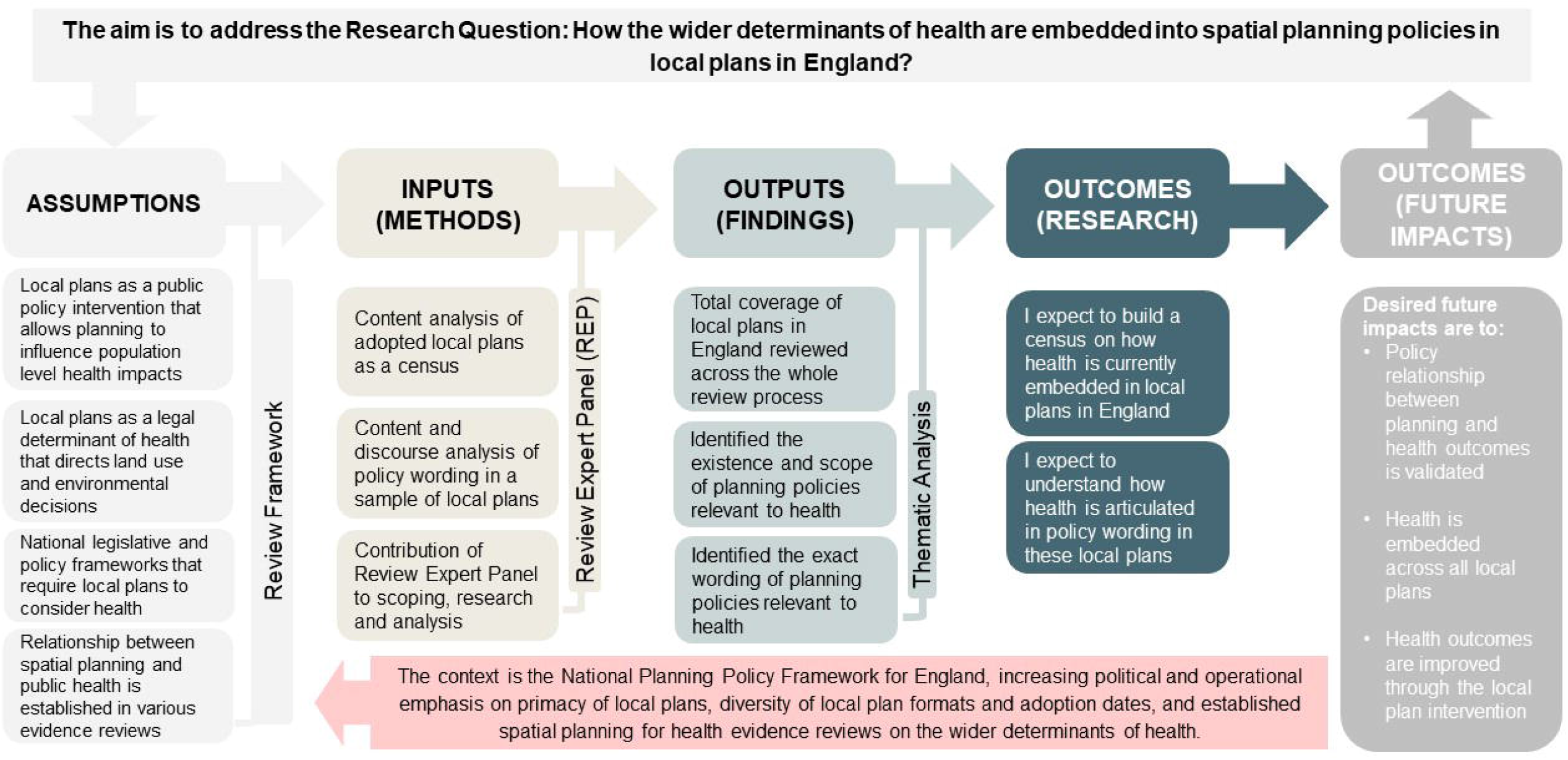
Early logic model for the local plans review

- To determine number of local plans with health policies that are policy compliant and/ or address all or some of the wider determinants of health.
- To understand how these health policies are worded/ phrased.

To have findings in presentable formats that are capable of comparative analysis to discuss and update relevant previous local plan reviews.

This protocol sets out a novel approach for a local plans review to gain a comprehensive understanding of the state of the current policy and practice, as the foundation for projecting our understanding of legal determinants of the health of future generations. The findings of the review are expected to have high policy relevance potential to national and local policy makers in the context of planning reform and increasing interest shown to use planning powers including through the loca plan to address public health priorities. The knowledge gained will be immensely beneficial to contribute to improving our awareness of local plans as a legal health determinant, to understand the scope of policy basis on which current planning decisions are being made on health, and whether this policy basis can be conducive to delivering on healthier environments in the future.

## 3. Methods and analysis

The review is based on a two-stage review:

- Stage 1 comprises of a local plans census. It is a desktop-based survey of each current local plan in England to be conducted between February and July 2023. In order to achieve the outcome of determining the scale of health policies present, the census will provide a complete enumeration of the ‘population’ of local plans at a point in time with respect to a set of defined health-oriented parameters.

- Stage 2 comprises of a local plans detailed review. It is a desktop-based content review on a selected sample of local plans from Stage 1 to be conducted between July and December 2023.It aims to allow a deeper dive to a finer granularity of the parameters based on qualitative content analysis of the policy texts.

A final stage, Stage 3, will form the next steps of the research in 2024-25 and involve a deep dive employing mixed methods review based on systems thinking including practical engagement with practitioners in local authority settings on their local plan preparation process.

### 3.1 Overall sample size

The sample pool will be based on the list of all local planning authorities (LPA) in England obtained from the official Department of Levelling Up, Housing and Communities planning application statistics on the gov.uk website. The review will note local government reorganisation in some areas which have combined many local authorities in two-tier areas into a single unitary authority. There are 338 LPAs which make up the total maximum sample pool.

### 3.2 A LPA is interpreted in primary legislation to include district councils

London borough councils, metropolitan district councils, county councils in relation to any area in England for which there is no district council, national park authorities and development corporations. The Planning Inspectorate keeps a Local Plan Tracker on all local plan status in all LPAs on its gov.uk website. The local plan document for each local authority will be the subject of the review. The local plan is also known as the core strategy or the development plan. It is the main document containing strategic policies but some local authorities will have another local plan that contains more detailed policies known as development management policies

### 3.3 Stage 1 census sampling framework with inclusion/ exclusion criteria

Stage 1 census inclusion/ exclusion criteria will include the following (see Figure 3 for full details of the stage 1 sampling framework:

**Figure 3.**
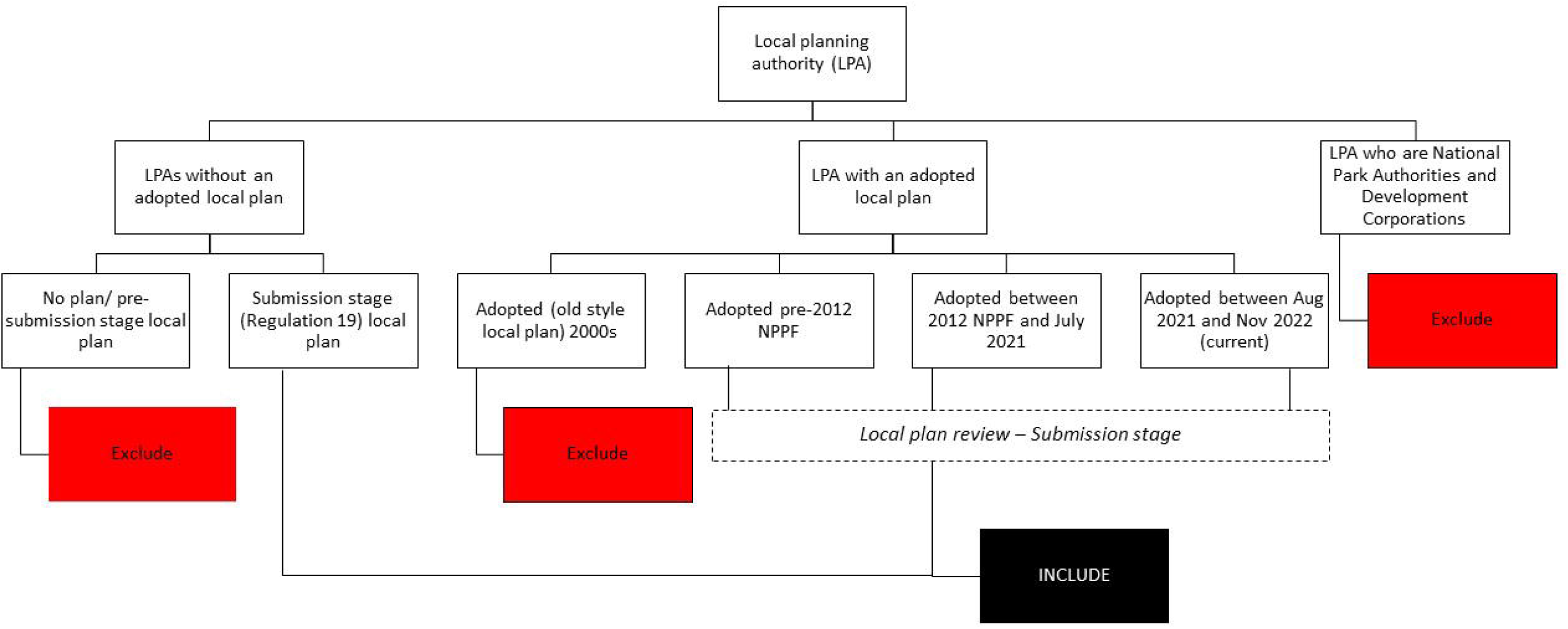
Stage 1 census sampling framework

- all LPAs with an ‘adopted’ local plan. An ‘adopted’ local plan is one that has been formally decided by the LPA to be accepted in accordance with relevant regulations. As of 2021, 90% of LPAs have an adopted local plan[17]. The author undertook a screening exercise in November 2022 in preparation for for Stage 1. It aimed to determine how many local plans would be eligible for inclusion in the review by using the latest PINS Local Plan Tracker data. The results of the exercise found 96% of LPAs have an adopted local plan so these plans will be included in the review.
- Many LPAs have commenced local plans reviews of their adopted local plans. Draft local plans at the Publication stage (a stage defined as an advanced stage in the plan-making process) will be included given recent case law about the material weight afforded to emerging local plans, and will be reviewed instead of the adopted plan they will ultimately replace.
- A small minority of LPAs that do not have an adopted local plan but have draft local plans at the Submission stage (Regulation 19) will be included.
- A small minority of LPAs have an old-style local plan adopted in the early 2000s but do not have a draft local plan in preparation will not be included.
- National Park Authorities are excluded because the expected scale of development and change is minor given their responsibility for natural areas of conservation.
- Development Corporations are excluded because they relate to a very small geographical area, and have responsibilities limited to development and regeneration and rely on the local plans of constituent local planning authorities.

### 3.4 Stage 2 detailed review sampling framework with further inclusion/ exclusion criteria Stage 2 detailed review will include further inclusion/ exclusion criteria to narrow down the sample size so that more comprehensive content analysis can be undertaken on a smaller number of local plans (see Figure 4 Stage 2 sampling framework). The criteria is

- Building on the inclusion/ exclusion criteria from Stage 1 census

**Figure 4.**
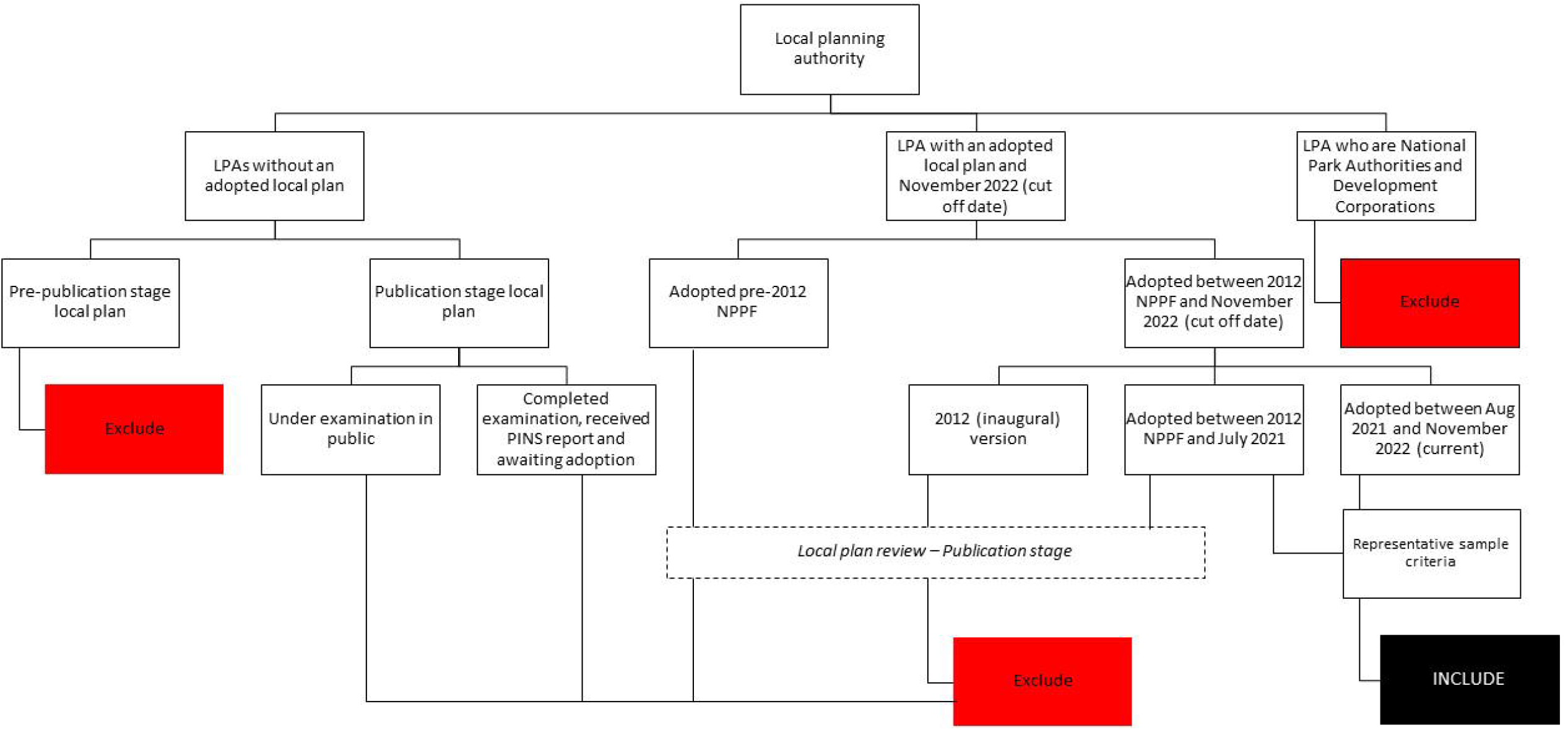
Stage 2 detailed review sampling framework
- Local plans that are ‘up-to-date’. An up-to-date plan is one that is adopted within 5 years of the adoption date for that plan. Planning regulations require local plans to be subject to review at least every five years. This additional criterion will cut the sample size for stage 2 further. As of 2022, 42% of LPAs have an adopted and up-to-date local plan[18].
- Representative sample will then be selected based on geography, local authority type and local plan adoption/ Publication year
- Representative sample will also be selected on their technical strength, ie responses within the local plan against each review parameter during the Stage 1 census review are positive against health.

### 3.5 Review framework

Table 1 sets out a summary review framework for the Stage 1 census which will be based on 16 parameter categories with sub-parameters to allow more details of the policy contents to be analysed for the Stage 2 detailed review. The full review framework with description and justification for each parameter and query function strategy is set out in Annex 1. Responses against each of the parameters will be obtained through 1) an interpretative word search of the whole local plan to determine the presence or not of health references, and the nature of this presence in actual policy wording or the supporting text to the policy, and 2) identification of policies in the table of contents of policies.

**Table 1:** Summary of the Local plans review framework for the Stage 1 census

| Main review parameters | Justification | Response (scoring) |
| --- | --- | --- |
| 1. Local Plan reference the statutory Joint Strategic Needs Assessment | NPPF policy compliance (paragraph 92 c) | ☐ Yes (1)<br>☐ No (0) |
| 2. Local Plan reference the statutory Joint Health and Wellbeing Strategy | NPPF policy compliance (paragraph 93 b) | ☐ Yes (1)<br>☐ No (0) |
| 3. Local Plan reference to other health-related strategy (for example healthcare facilities, obesity or healthy weight) | NPPF policy compliance (paragraph 93 b) | ☐ Yes (1)<br>☐ No (0) |
| 4. Local Plan subject to a standalone health impact assessment | Upstream health effects of plan assessed. | ☐ Yes (1)<br>☐ No (0) |
| 5. Local Plan strategic objective for health | Local authority spatial aim | ☐ Yes (1)<br>☐ No (0) |
| 6. Local Plan strategic policy on health | Local authority health policy | ☐ Yes (1)<br>☐ No (0) |
| 7. Local Plan policy on Housing:<br>a) affordable housing<br>b) energy efficient homes<br>c) housing to meet older people and other special needs | Evidence-informed policy [7] | For each parameter:<br>☐ Yes (reference to health in policy) (3)<br>☐ Yes (reference in supporting text)(2)<br>☐ Yes (but no reference to health)(1)<br>☐ No (0) |
| 8. Local Plan policy on Neighbourhood Design:<br>a) design quality<br>b) compact neighbourhood/ access to services |  |  |
| 9. Local Plan policy on transport: active travel |  |  |
| 10. Local Plan policy on Natural and Sustainable Environment:<br>a) green spaces<br>b) flood risk management<br>c) urban heat island and overheating<br>d) air and noise pollution |  |  |
| 11. Local Plan policy on Food Environment:<br>a) hot food takeaways<br>b) food growing |  |  |
| 12. Local Plan policy on social value and use of developer contributions on social infrastructure | Industry-informed policy and practice to meet social responsibility aim |  |
| 13. Local Plan policy on use of health impact assessment in planning applications | Planning Guidance compliance | ☐ Yes (with trigger)(3)<br>☐ Yes (no trigger)(2)<br>☐ Yes (supporting text) (1)<br>☐ No (0) |
| 14. Local Plan policy on use of key national healthy planning frameworks or accreditation schemes | Industry-informed policy on voluntary frameworks | ☐ Yes (reference to health) (3)<br>☐ Yes (reference in supporting text)(2)<br>☐ Yes (but no reference to health)(1)<br>☐ No (0) |
| 15. Local Plan monitoring indicators relevant to health under Parameters 7-13 | Evidence-informed process to measure progress | ☐ Yes (1)<br>☐ No (0) |
| 16. Supplementary indicators to reflect current public health policy interests in planning, ie gender inequalities, mental wellbeing, crime prevention, and suicide prevention. | Evidence-informed process to measure progress on areas of interest | ☐ Yes (1)<br>☐ No (0) |

The aim is to determine the presence or absence of health references, and the nature of this presence in actual policy wording or the supporting text to the policy. The word search is interpretative because the absolute presence of health references may not always indicate consideration of health. Therefore once the words are searched and identified, a process of review of context policy wording will be undertaken to determine the response.

This review framework and its parameters is justified by seeking to address current policy compliance requirements in the NPPF in relation to promoting healthy and safe communities, and contemporary areas of interest expressed by policy makers and practitioners[9, 19].

The rest of the parameters are justified by being underpinned by published evidence, namely the evidence themes presented in Spatial Planning for Health: An evidence resource for planning and designing healthier places published by Public Health England with the underlying research subjected to an umbrella review undertaken by the University of the West of England [6, 7]. Figure 1 indicates what and how specific planning principles and modifiable features can impact on health outcomes. It was not the aim of the research to conduct a systematic review (of which there are several recent reviews (see for example [20]) to seek further validation of what is already known or that similar themes have consistently emerged from such reviews (see for example [21]). Therefore the adoption of PHE’s evidence resource allows the research to progress to determining how health determinants are currently embedded into policy and practice while acknowledging the potential for future evidence to indicate emerging determinants.

It is not the purpose of the Stage 1 census to assess the quality and effectiveness of the policy but the presence and explicit reference to health. Scoring will be attributed to each response for the purpose of listing review results and ability to conduct quantitative analysis to the responses but not for ranking. The responses will be binary which means there will be no more than one positive response per parameter. For example, if there are requirements for health in a specific policy as well as in the supporting text, a positive response will only be attributed to the former as that will be the maximum and preferred response.

A test and trial phase was undertaken to test and refine the review framework during December 2022-January 2023 on two sample local plan before commencing the full review. No analysis and synthesis took place yet at this stage of the research. The main observations of the review framework from this test phase:

- indicated the need to set out clear and unambiguous key words to ensure the proper identification of contents while providing the flexibility and opportunity during the search to identify associated and synergistic contextual contents.
- suggested key word searches still require an element of interpretation and checking depending on where these words were identified in the document.
- questioned employing NViVO during the study. The use of automated query functions on NViVo was initially assumed to be most efficient for the Stage 1 census given the large number of documents involved. But the author observed using NViVo requires an additional step in the workflow, ie collation of local plan documents, which do not add material value to the review, because interpretative review of identified policies is required anyway. So the use of NViVo would be more appropriate with smaller sample of local plans in Stage 2 and maximise its functionality over basic PDF.
- given the above, it found that centralising the collection of review data on one platform such as on Excel can provide a sound basis for ensuring consistency and oversight, and ultimately analysis and presentation of results.

As a result of the test and trial phase the review framework and data collection process will be amended with main search terms added to the review framework as a guide to identifying and collecting relevant text (annex 1), and supplementing term searches with searches of the table of contents or list of policies section of the local plan to find relevant policies.

### 3.6 Data analysis plan

The data collection tools for each local plan review for stages 1 and 2 will be undertaken using a combination of methods. Stage 1 will be primarily undertaken through key word searches of PDF documents while data extraction of relevant policy wording will also be undertaken. Data presentation will be collated on Excel as the master document. Using the data extracted, Stage 2 will introduce the use a data management and Excel to centralise the collation of, refine, analyse and present results.

Adopting an appropriate method of analysis following the review has taken place is dependent on what and how raw primary data will be collected. Stage 1 census will predominantly adopt the Content method with deductive reasoning to state whether the local plan contains X or Y with secondary variables that can be quantified. The data collection method will be a simple response-collection system based on binary answers without weighting as proposed in Table 1. Stage 2 will predominantly adopt a Discourse method to identify the exact policy wording adopted and to review the language used in these policy wording as a sign of strength or weakness of action against health outcomes.

The data will be able to be presented in quantitative and qualitative formats depending on the audience and purpose of dissemination. Multiple presentation styles will be used such as tables or graphs, as well as the ability to present responses according to spatial, geographical and temporal associations, such as through geographical and systems maps, and timelines based on local government area characteristics such as deprivation and urban/ rural classification [22].

There is a variety of methods and in practice the final method employed will be a combination of methods given the comparative nature between some of the methods, ie content, thematic and thematic analyses. This is because it is unlikely any analysis will be conducted without elements of framework analysis using a preconceived framework of parameters to adhere to, because without it, the analysis process would be aimless. In general, the overall documentary analysis method for the Stage 1 census will have a quantitative inductive content analysis[23].Stage 2 detailed review will comprise of the phases of thematic analysis[24, 25] most commonly associated with the grounded theory[26, 27]. While data will be structured according to review parameters, it is not known at the outset how they will be set out in local plans which provides the opportunity to apply the grounded theory process to be guided by what the data will reveal. The author is the primary investigator of data collection and analysis and follows a defined and systematic review framework as set out in Table 1 in order to induct themes from the data. This particular school of thought in constructivist grounded theory [28] is an appropriate method to employ and aligns well with the discretionary nature of decision-making in the UK planning system.

### 3.7 Expert involvement through the Review Expert Panel

For the purpose of ensuring the robustness and relevance of the research activity a Review Expert Panel has been established. A Review Expert Panel will act in a similar capacity of Patient and Public Involvement but with engagement with experts. The expert members of the panel are: associate director in a private sector planning consultancy, managing director of a healthy building consultancy, head of a planning improvement service, planning for health specialists in local government, a public health lecturer and a systems lecturer. There are no patients or members of the public involved in the Panel. The Panel will help shape and review the protocol and progress; and commenting on and contributing insight on research, policy and practice implications from research findings. A first meeting of the Panel was held in December 2022 in which the Terms of Reference was discussed and agreed, and respective participation consent from individuals were received.

### 3.8 Strengths and limitations of the study

- The study is novel because local plan reviews are not commonly done given the scale of the sampling pool so this study will be novel and seminal as a baseline study.
- Local plans are an ideal source of data as they are static public policy documents with shared national policy requirements which make them ideal for content and thematic analysis.
- The review process is robust because it is sense checked through the systematic involvement and contribution of a panel of subject matter experts.
- Sampling strategy is limited because it does not completely reflect the complexity of different local plan-making timescales and formats with many local plans subject to regular review. So this makes direct comparisons challenging.
- Data identification and coding framework of review parameters is limited and dependent on professional interpretation of local plans so the data is depending on the prior experience/ qualification of the researcher.

### 3.9 Patient and public involvement

None.

## Supporting information

10.Annex 1: Details of the review framework

## Data Availability

All data produced in the present study are available upon reasonable request to the authors.

## 4. Ethics and dissemination

The local plans review is desktop based using publicly available policy document so ethics approval will not be required. Any primary and synthesis data collected during the review will be stored securely according to university regulations. For the Review Expert Panel, ethics approval is not needed as determined by an initial checklist application to the University’s Research Ethics Committee.

Results of the study will be presented in public health and professional development conferences and subject to submission to international planning and public health journals. Briefings will also be produced for policy makers and professional audiences working in national and local government.

## 5. Authors’ contributions

MC – conceptualisation, data curation, formal analysis, investigation, methodology, writing, project administration, agree to be accountable as author. NC – supervision, methodology, review writing, agree to be accountable as author

## 6. Reporting Guidelines

Repository: Chang, Michael (2023): SRQR_checklist_FINAL.pdf. figshare. Online resource. https://doi.org/10.6084/m9.figshare.22210405. Data are available under the terms of Creative Commons CC BY 4.0.

## 9. Data (and Software) Availability

No data are associated with this article.

## 10. Annex 1: Details of the review framework

## Footnotes

1 Note that OHID replaced Public Health England’s health improvement functions from October 2022.

