## Supplementary material for "Review of spatial planning policy as an intervention for improving public health: protocol for a health-focused content review of local plans in England": 10.Annex 1: Details of the review framework

| Parameters | Description | Key search terms/ locations |
|---|---|---|
| Basic identifier data such as: administrative English region, LA type classification, and date of local plan adoption | Providing the baseline data about the LPA and the timelines of preparing the local plan, to provide comparative characteristics for later analysis. Further data points can be included during analysis such as deprivation levels etc. | Not applicable |
| 1. Does the Local Plan reference the Joint Strategic Needs Assessment? | These parameters will provide an understanding of policy compliance with the NPPF for England which sets out the land use policies and expectations of the Government.<br><br>In particularly to identify how each local plan address existing NPPF requirements for planning policies to consider a. identified health and well being needs, ie the joint strategic needs assessment, and b. help support the delivery of health and wellbeing strategies, ie the joint health and wellbeing strategy. These requirements have been in place since the first NPPF was published in 2012. Each local authority with public health responsibilities are required to create a joint strategic needs assessment and a joint health and wellbeing strategy. | 'joint strategic needs assessment', 'JSNA', 'health needs', 'wellbeing needs' |
| 2. Does the Local Plan reference the Joint Health and Wellbeing Strategy? | | 'health and wellbeing strategy', 'joint health', 'joint local health' |
| 3. Does the Local Plan reference any other health-related strategy (for example healthcare facilities, obesity and healthy weight)? | | 'obesity strategy', 'estates strategy', 'health strategy' |
| 4. Was the Local Plan subject to a standalone health impact assessment? | Providing an understanding of whether and how health considerations have been integrated into the plan-making process as the sustainability appraisal is a legal requirement where population and human health is a legal factor to be assessed. Although a separate HIA is not required. | 'health impact assessment', 'HIA' |
| 5. Does the Local Plan have a strategic objective for health? | Understand if local plans have health as a spatial objective and policy as an indicator of strategic commitment to health. Often local plans refer to corporate plans or local community plans which usually have healthy places-related objectives. The specific wording of the policy will be identified. | Vision and Objectives section of the local plan |
| 6. Does the Local Plan have a strategic policy on health? | | Table of contents or list of policies |
| 7. Does the Local Plan have a policy on Housing:<br>a) Affordable housing<br>b) energy efficient homes<br>c) housing to meet older people and other special needs | In addition to cross referencing to relevant requirements in the NPPF, these parameters are based on the PHE evidence review's themes (Public Health England, 2017) which are based on literature review undertaken by the University of the West of England (Bird et al., 2018). Questions on each theme will be broken down by Planning Principles to provide deeper breakdown and nuance in the wider determinants. | 'affordable housing' or 'healthy homes' or 'homes for specific needs' 'older people housing' or 'specialist or supported housing' or 'housing quality' or 'energy efficient homes' |
| 8. Does the Local Plan have a policy on Neighbourhood Design: | These parameters allow identification of whether there are policies that include the health determinants with evidence suggesting leading to positive health | 'compact neighbourhoods' or 'walkable neighbourhoods' or 'good design' or 'urban design' or 'access to services' |

| | | |
|---|---|---|
| a) design quality<br>b) compact neighbourhood/ access to services | outcomes, and collection of a policy wording that can form a database of examples. | |
| 9. Does the Local Plan have a policy on Transport: active travel, walking and cycling | | ‘active travel’ or ‘walking and cycling’ |
| 10. Does the Local Plan have a policy on Natural and Sustainable Environment:<br>a) green spaces<br>b) flood risk management<br>c) urban heat island and overheating<br>d) air and noise pollution | | ‘greenspaces’ or ‘natural environment’ or ‘flood risk management’ or ‘sustainable urban drainage/ SUDs’ or ‘waterways’ or ‘green/ blue infrastructure’ or ‘air pollution’/ ‘air quality’ or ‘noise pollution’/ ‘noise quality’ |
| 11. Does the Local Plan have a policy on the Food Environment:<br>a) hot food takeaways<br>b) food growing | | ‘hot food takeaways’ or ‘food environment’ or ‘fastfood’ or ‘food growing’ or ‘community gardens’ or ‘allotments’ or ‘urban farming’ |
| 12. Does the Local Plan have a policy on social value and use of developer contributions on social infrastructure? | Social value in the context of the built environment is when buildings, places and infrastructure support environmental, economic and social wellbeing. While the social value duty on public bodies is not specific to spatial planning and applies to procurement, it would be interesting to see how and whether any LPAs have identified opportunities of using the local plan as a lever to deliver social value in the built environment (UK Green Building Council, 2020). | ‘social value’ or ‘community infrastructure’ or ‘social infrastructure’ or ‘community facilities’ |
| 13. Does the Local Plan have a policy on requiring health impact assessment in planning applications? | Providing an understanding of awareness of national planning guidance that enables local requirement for planning applications to submit a HIA where there are expected to be significant impact. | ‘health impact assessment’, ‘HIA’ |
| 14. Does the Local Plan require the use of key national healthy planning frameworks or accreditation schemes? | Providing an identification of examples of local innovation if other health (direct/ indirect) tools are required or referenced in policy or supporting text. | ‘Building for a Healthy Life’ or ‘Building for Life’ or ‘lifetime homes/ neighbourhoods’ or ‘Home Quality Mark/ HQM’ or ‘Code for Sustainable Homes’ or ‘active design’ or ‘Planning for Health’ |
| 15. Does the Local Plan have monitoring indicators relevant to health under | Providing an important indication of what and how LPAs will evaluate the outcomes of implementing their policies on health, as legally required to be reported in annual Authority Monitoring Reports. UK | Monitoring Framework section of the local plan |

| | | |
|---|---|---|
| each of the Parameters 7 -13? | government has not established and required a set of national indicators and it is up to LPAs to create and monitor against any indicator as part of the AMR process. | |
| 16. Supplementary: Does the Local Plan include policy considerations on:<br>a) <u>gender inequalities</u><br>b) <u>mental wellbeing</u><br>c) <u>crime prevention</u><br>d) <u>Suicide prevention</u> | Providing an important high level recognition of emerging political, policy, practice and research priorities on aspects of planning, built and natural environments in relation to health inequalities. | ‘gender’ or ‘mental’ or ‘crime’ or ‘suicide’ |
